# Hypothalamic integrity is associated with age, sex and cognitive function across lifespan: A comparative analysis of two large population-based cohort studies

**DOI:** 10.1101/2024.04.29.24306537

**Authors:** Peng Xu, Santiago Estrada, Rika Etteldorf, Dan Liu, Mohammad Shahid, Weiyi Zeng, Deborah Früh, Martin Reuter, Monique M.B. Breteler, N. Ahmad Aziz

**Affiliations:** Population Health Sciences, German Center for Neurodegenerative Diseases (DZNE), Bonn, Germany; Artificial Intelligence in Medical Imaging, German Center for Neurodegenerative diseases (DZNE), Bonn, Germany; A.A. Martinos Center for Biomedical Imaging, Massachusetts General Hospital, Boston MA, USA; Department of Radiology, Harvard Medical School, Boston MA, USA; Institute for Medical Biometry, Informatics and Epidemiology (IMBIE), Faculty of Medicine, University of Bonn, Germany; Department of Neurology, Faculty of Medicine, University of Bonn, Germany

**Author notes:** **Corresponding author:** Dr. N. Ahmad Aziz, MD PhD, Population Health Sciences, German Center for Neurodegenerative Diseases (DZNE), Venusberg-Campus 1/99, 53127 Bonn Germany. **Contributors:** PX, MMBB, and NAA conceptualized the project. The first draft of the manuscript was written by PX and NAA. Data were analyzed by PX, SE, RE, DL, WZ, DF, and NAA. SE, MS, MR, and MMBB provided technical, statistical and methodological advice. All authors provided critical feedback and contributed to the writing and revision of the final version of the manuscript. **Data availablility statement:** The Rhineland Study’s dataset is not publicly available because of data protection regulations. Access to data can be provided to scientists in accordance with the Rhineland Study’s Data Use and Access Policy. Requests for further information or to access the Rhineland Study’s dataset should be directed to. All individual level data from the UK Biobank Imaging Study used in the current manuscript are available through the UK Biobank Resource (https://www.ukbiobank.ac.uk). This research has been conducted using the UK Biobank Resource under Application Number 82056.

## Abstract

**Background:** The hypothalamus is the body’s principal homeostatic center. Emerging findings from animal studies suggest that the hypothalamus could also play a crucial role in the modulation of cognition. However, detailed assessments of age and sex effects on hypothalamic structural integrity and its cognitive correlates across the lifespan are still lacking. Therefore, we aimed to investigate hypothalamic structural integrity in relation to age, sex and cognitive performance across lifespan in the general population.

**Methods:** We used cross-sectional data from the Rhineland Study (RS) (N=5812, 55.2 ± 13.6 years, 58% women) and the UK Biobank Imaging Study (UKB) (N=45076, 64.2 ± 7.7 years, 53% women), two large-scale population-based cohort studies. Volumes of hypothalamic structures were obtained from 3T structural magnetic resonance images through application of a recently developed automatic parcellation procedure (FastSurfer-HypVINN). The standardized cognitive domain scores were derived from extensive neuropsychological test batteries. We employed multivariable linear regressions to assess age and sex effects on volumes of hypothalamic structures, and to evaluate the associations of these volumes with domain-specific cognitive function.

**Findings:** Mean (standard deviation) volumes of the total hypothalamus were 1124.2 mm^3^ (104.8) in RS and 1102.1 mm^3^ (119.9) in UKB. With increasing age, the volumes of the total, anterior and posterior hypothalamus, and mammillary bodies decreased (between -1.20 to -0.14 mm^3^/year in RS and between -3.82 to -0.49 mm^3^/year in UKB), and of the medial hypothalamus and tuberal region increased (between 0.33 to 0.65 mm^3^/year in RS and between 0.21 to 0.68 mm^3^/year in UKB). Volumes of all hypothalamic structures were larger in men compared to women. Larger total hypothalamus volumes were associated with better global cognition (β ± standard error (SE): 0.025 ± 0.017 [RS] and 0.026 ± 0.007 [UKB], both p<0.005), and total memory (0.030 ± 0.022 [RS] and 0.021 ± 0.009 [UKB], both p<0.007), while larger posterior hypothalamus volumes were associated with better global cognition (0.036 ± 0.014 [RS] and 0.028 ± 0.006 [UKB], both p<0.001), and total memory (0.038 ± 0.018 [RS] and 0.020 ± 0.008 [UKB], both p<0·001).

**Conclusion:** We found strong age and sex effects on hypothalamic structures, as well as robust associations between these structures and domain-specific cognitive functions. Overall, these findings thus implicate specific hypothalamic subregions as potential therapeutic targets against age-associated cognitive decline.

## Introduction

The human hypothalamus, located beneath the thalamus, is part of the diencephalon.^1^ It has a complex and delicate architecture, consisting of dozens of interconnected nuclei and tracts, and plays an essential role in the regulation of a wide range of physiological, behavioural, and cognitive processes.^2-4^ Moreover, accumulating neuropathological evidence points towards extensive hypothalamic involvement in various neurodegenerative disorders, including Alzheimer disease,^5^ Parkinson disease,^6^ Huntington disease,^7^ frontotemporal dementia,^8^ and amyotrophic lateral sclerosis.^9^ Assessing hypothalamic structure and its phenotypic correlates across the lifespan could therefore provide novel insights into the neurobiological basis of age-associated dysregulation of a range of bodily functions, particularly those occurring in the context of age-related neurodegenerative diseases.^5-9^

Neuropathological studies have substantially advanced our understanding of the functional anatomy of different hypothalamic structures in humans^.4,10^,11 However, limited sample sizes, peri- and postmortem factors affecting brain tissue quality, application of a range of different neuropathological techniques, as well as limited availability of ante-mortem clinical data pose major challenges that remain to be overcome. Therefore, structural magnetic resonance imaging (MRI) studies have been increasingly employed to assess hypothalamic structural integrity *in vivo*.^12-15^ While these studies have provided preliminary estimates of age and sex effects on hypothalamic volume, the results have been inconsistent, likely due to inclusion of limited age ranges, small sample sizes, and pre-selection of specific (diseased) populations. Hence, more in-depth investigations of the structural integrity of the hypothalamus in relation to age and sex in much larger populations are warranted for a better understanding of the role of this tiny, yet crucial brain structure in health and disease.

Cognitive impairment is the hallmark of many age-associated neurodegenerative diseases.^16^ Recent findings from animal experiments point towards a crucial role of the hypothalamus in the modulation of cognition, both directly through extensive projections to cortical and limbic regions and indirectly through regulation of mood, sleep/circadian rhythm, metabolism, and neuroendocrine outflow^.2-4,12^,17 For example, as an important subregion of the hypothalamus and a key component of the circuit of Papez, the mammillary bodies are crucially implicated in the regulation of cognition, especially memory function, through their extensive connections to the hippocampus and other cortico-limbic structures.^18^ However, despite consistent evidence from animal experiments for the critical role of the mammillary bodies for cognitive performance, results of human studies assessing the association between volumes of mammillary bodies and cognitive impairment are inconsistent.^19,20^ Similarly, recent animal experiments indicate that disturbances of the hypothalamic orexinergic system could be implicated in the pathogenesis of both mild cognitive impairment and Alzheimer disease, potentially through modulation of amyloid-β dynamics.^21,22^ Moreover, hyperactivation of the hypothalamic-pituitary-adrenal axis, which is under direct control of the corticotropin releasing factor-producing neurons of the hypothalamic paraventricular nucleus (PVN), has for long been implicated in the pathogenesis of age-associated cognitive decline.^23,24^ Mounting evidence indicates that also other hypothalamic substructures could be related to cognitive function, including the lateral hypothalamus, the infundibular nucleus, and the supra-mammillary nucleus.^2,25^ Nonetheless, the relevance of these findings for age-associated cognitive decline and neurodegenerative diseases in humans remains unclear as the association between hypothalamic structure and cognitive function has not been systematically assessed before in large well-characterized (population-based) cohorts, mainly due to lack of fully automatic hypothalamic parcellation procedures required for large-scale deployment.

To enable detailed assessment of the human hypothalamus in large cohorts, recently we developed a fully automatic, multi-modal parcellation procedure for accurate volumetric segmentation of the hypothalamus and its subregions on brain MRI.^26^ Here, to address the aforementioned issues, we applied this automatic segmentation algorithm to the full brain imaging datasets of two independent large-scale population-based studies for cross-validation, i.e. the Rhineland Study and the UK Biobank Imaging Study. Specifically, we aimed to investigate the structural integrity of the human hypothalamus and its subregions in relation to age, sex, as well as detailed domain-specific measures of cognitive performance in the general population. Our findings indicate that the hypothalamus undergoes profound region-specific changes across lifespan, exhibits substantial sex differences and is strongly related to both general and domain-specific cognitive function.

## Methods

### Study cohorts

The Rhineland Study (RS) is an ongoing population-based cohort study, recruiting inhabitants aged 30 years and above from two geographically defined areas in Bonn, Germany (www.rheinland-studie.de). The study’s only exclusion criterion is an insufficient command of the German language required for providing informed consent. Participants are primarily Caucasians of European descent.^27^ The study aims to contribute to the prevention, early detection and treatment of neurodegenerative and other age-related diseases. Participants undergo an ∼8-hour deep phenotyping assessment, including detailed brain imaging and standardized cognitive test batteries. The study protocol was approved by the ethics committee of the University of Bonn Medical Faculty (Ref: 338/15). The study is conducted according to the International Conference on Harmonization Good Clinical Practice standards (ICH-GCP), with written informed consent obtained in accordance with the Declaration of Helsinki.

The UK Biobank Imaging Study (UKB) is part of the UK Biobank Study, an ongoing population-based cohort study collecting in-depth genetic and health information data on approximately half a million participants from across the United Kingdom. In 2014, the UK Biobank Study re-invited participants who had completed the baseline visit for an additional visit, involving detailed brain imaging and cognitive testing batteries. The selection of participation for the this sub-study was based on protocols of the UK Biobank imaging enhancement.^28^ Approval for conducting the study was received from the National Information Governance Board for Health and Social Care and the National Health Service North West Centre for Research Ethics Committee (Ref: 11/NW/0382). Every participant provided written informed consent.

Our initial study population consisted of the first 8318 participants of the RS, and the 49304 participants of the UKB who had completed the first imaging visit. We excluded 2180 individuals from the RS without brain imaging data. Subsequently, after visual quality control assessments, we excluded 326 individuals from the RS dataset and 4228 individuals from the UKB dataset due to insufficient image quality. This resulted in a final analytical sample size of 5812 and 45076 participants from the RS (**Supplementary Figure 1**) and UKB cohorts (**Supplementary Figure 2**), respectively.

### Imaging data

In RS, imaging data were obtained using two identical 3T MRI scanners equipped with 64-channel head-neck coils (MAGNETOM Prisma; Siemens Healthcare) at two examination facilities situated in Bonn, Germany.^29^ Detailed information about the protocols and sequence parameters in RS are provided in **Supplementary Table 1**.

MRI data in UKB were obtained from Siemens Skyra 3T scanners equipped with 32-channel head coils. Initially, a single scanner in Cheadle Manchester was dedicated to the UKB, followed by two further identical scanners in Newcastle and Reading.^30^ The complete protocols and sequence parameters in UKB are available in the online documentation (biobank.ctsu.ox.ac.uk/crystal/crystal/docs/brain_mri.pdf).

We applied our automated deep learning-based method ‘FastSurfer-HypVINN’ for sub-segmentation of the hypothalamus on 0.8 mm isotropic T1- and T2-weighted images from RS, as well as 1.0 isotropic T1-weighted images from UKB. Importantly, we previously validated the performance of FastSurfer-HypVINN against ground-truth manual segmentations on MR images from both RS and UKB.^26^ This pipeline segments the hypothalamus into six subregions on each side, including the anterior hypothalamus (AH), the medial hypothalamus (MH), the lateral hypothalamus (LH), the tuberal region (TBR), the posterior hypothalamus (PH), and the mammillary bodies (MMB).^26^ Details about the neuroradiological definition of different hypothalamic structures are provided in **Supplementary Table 2**. Finally, total brain volume and estimated total intracranial volume (ETIV) for both datasets were obtained using the FreeSurfer (version 6.0) standard processing pipeline.^31,32^

### Demographic and cognitive data

Age and sex were based on self-reports. Level of education was assessed according to the International Standard Classification of Education 2011 (ISCED) (RS), or an educational attainment score based on the highest qualification level (UKB) as described previously.^33,34^ In RS, four fluid cognitive domains were evaluated through multiple tests: working memory (Corsi Block-tapping Test and Digit Span Test), episodic verbal memory (Auditory Verbal Learning and Memory Test (AVLT)), executive function (Word Fluency Task, Trail-Making Test B, and Anti-saccade Task), and processing speed (Trail-Making Test A and Pro-saccade Task).^35^ Similarly, UKB employed seven tests for the assessment of the same cognitive domains: working memory (Numeric Memory Test), episodic verbal memory (Paired Associate Learning Test and Pairs Matching Test), executive function (Trail-Making Test B), and processing speed (Trail-Making Test A, Reaction Time Test, Symbol Digit Substitution Test).^36^ Detailed information regarding all cognitive tests included in the present study are provided in **Supplementary Table 3**.

### Cortisol levels

Cortisol levels were only available in a subset of participants of the Rhineland Study (N=1515). Cortisol concentrations were quantified in scalp hair samples through liquid chromatography tandem mass spectrometry using a steroid panel as described previously.^37^ Scalp hair cortisol levels reflect the average amount of cortisol (in pg/mg) in the three months prior to the sampling date.

### Statistical analysis

Results are reported as mean ± standard deviation (SD) for continuous variables, and as counts and percentages for categorical variables, unless otherwise specified.

#### Age and sex

We used multivariable linear regression analyses to assess the effects of age and sex (independent variables) on volumes of the hypothalamus and its subregions (dependent variables). In these models, we also included a quadratic term for age to evaluate potential nonlinear age effects, and a product term between age and sex to evaluate potential age × sex interaction effects, while adjusting for ETIV to account for differences in head/brain size. All continuous variables were mean-centred to mitigate collinearity. In the event that the higher-order or interaction terms were not statistically significant, these terms were removed from the final models to obtain the most precise estimates for the main effects. For bilateral hypothalamic structures, we used the sum of the left and right sides as the outcome for conciseness, given that initial analyses per side did not reveal any lateralized effects (data not shown).

#### Cognition

Cognitive test scores with skewed distributions were log-transformed and, if applicable, inverted to ensure that higher values represented better cognitive performance. Subsequently, all cognitive test scores were Z-standardized to enable direct comparison of the effect sizes across multiple cognitive domains. To produce summary cognitive domain scores, we averaged the corresponding Z-scores of tests belonging to each cognitive domain (**Supplementary Table 3**). Additionally, we averaged the domain scores of working memory and episodic verbal memory to produce the domain score for total memory. For producing an overall global cognition score, we averaged the Z-scores across all domains. As a sensitivity analysis, we also created an overall global cognition score using the first principal component of the Z-scores across all cognitive tests, which accounted for 34.7% (RS) and 34.8% (UKB) of the total variance in cognitive performance, as a summary measure (**Supplementary Figure 3**). Multivariable regression models were employed to evaluate the association between the volumes of different hypothalamic structures (independent variables) and cognitive domain scores (dependent variables), while adjusting for age, age^2^, sex, education level, and ETIV. All effect sizes are reported as standardized effect sizes, i.e., indicating a change in SD of cognitive domain scores per SD change of volume.

#### Cortisol

For direct functional validation of a part of our findings, we assessed the association between the volume of MH, which includes the corticotropin releasing factor-producing neurons of the PVN, and cortisol levels. To this end, we employed multivariable regression models with age, age^2^, sex, MH volume (adjusted for ETIV) and its interaction with age as independent variables, and log-transformed cortisol levels (to account for the right skewed distribution of hair cortisol levels) as the dependent variable.

All models were run in the two cohorts (i.e., RS and UKB) separately, including additional sensitivity analyses in which we excluded participants with known neurological diseases (including stroke, dementia, Parkinson disease, and multiple sclerosis). Statistical analyses were performed in R (base version 4.2.2). Two-sided p-values < 0.05 were considered statistically significant. As comparisons of all findings between two large independent cohorts already ensured direct assessment of their external validity, we refrained from additional multiple comparisons corrections.

## Results

### Demographics

The mean age of the study participants was 55.2 ± 13.6 years (58% women) in RS, and 64.2 years ± 7.7 (53% women) in UKB. Mean volumes of the total hypothalamus (TH) were 1124.2 ± 104.8 mm^3^ in RS and 1102.1 ± 119.9 mm^3^ in UKB (**Table 1**). On average, excluded participants were older, more often male, and had a lower education level compared to those included in the analytical sample (RS: **Supplementary Table 4**, UKB: **Supplemental Table 5**).

### Age and sex effects on hypothalamic volumes

With increasing age, the volumes of TH, AH, PH and MMB decreased (with between -1.20 to -0.14 mm^3^/year in RS, and between -3.82 to -0.49 mm^3^/year in UKB), while the volumes of MH and TBR increased (with between 0.33 to 0.65 mm^3^/year in RS, and between 0.21 to 0.68 mm^3^/year in UKB) (**Table 2** and **Figure 1**). The association between age and LH volume was weaker and inconsistent in the two datasets, with LH volume exhibiting an increase with age in RS and a decrease with age in UKB (**Table 2**).

**Table 1.** Characteristics of the analytical sample from the Rhineland Study and the UK Biobank Imaging Study.

|  | Rhineland Study<br>N = 5812 | UK Biobank Imaging Study<br>N = 45076 | P-value |
| --- | --- | --- | --- |
| Age, years | 55.2 (13.6) | 64.2 (7.7) | <0.001<sup>a</sup> |
| Age range, years | 30 - 95 | 44 - 83 |  |
| Sex |  |  | <0.001<sup>a</sup> |
| Women | 3,362 (58%) | 23,950 (53%) |  |
| Men | 2,450 (42%) | 21,126 (47%) |  |
| Education level<sup>b</sup> |  |  | <0.001<sup>c</sup> |
| High | 3,793 (66%) | 21,811(49%) |  |
| Middle | 1,869 (32%) | 19,935 (45%) |  |
| Low | 95 (1.7%) | 2,855 (6.4%) |  |
| Neurological diseases<sup>d</sup> | 113 (1.9%) | 746 (1.7%) | <0.001<sup>e</sup> |
| ETIV, ml | 1,548.8 (147.8) | 1,466.7 (158.7) | <0.001<sup>f</sup> |
| TBV, ml | 1,103.6 (117.7) | 1,108.6 (107.5) | <0.001<sup>g</sup> |
| Volumes, mm<sup>3</sup> |  |  |  |
| Total Hypothalamus | 1,124.2 (104.8) | 1,102.1 (119.9) | <0.001<sup>g</sup> |
| Anterior Hypothalamus | 272.3 (46.4) | 221.0 (51.5) | <0.001<sup>g</sup> |
| Medial Hypothalamus | 104.0 (26.0) | 111.0 (29.8) | <0.001<sup>g</sup> |
| Tuberal Region | 43.3 (9.6) | 44.7 (10.0) | 0.57<sup>g</sup> |
| Lateral Hypothalamus | 135.6 (21.3) | 139.8 (25.4) | <0.001<sup>g</sup> |
| Posterior Hypothalamus | 368.4 (45.3) | 368.9 (57.3) | <0.001<sup>g</sup> |
| Mammillary Bodies | 209.5 (26.7) | 216.7 (29.8) | <0.001<sup>g</sup> |

Data are shown as mean (SD) or n (%) for continuous and categorical variables, respectively.
<sup>a</sup>) P-values were obtained from Welch's Two Sample t-test for continuous variables and from Pearson's Chi-squared test for categorical variables.
<sup>b</sup>) Education level in the Rhineland Study was defined as follows: High (ISCED-11 levels 6-8), Middle (ISCED-11 levels 3-5), and Low (ISCED-11 levels 0-2). In the UK Biobank Imaging Study, it was classified as: High (equal to or higher than college level, Middle (secondary education level), and Low (below secondary level). These two classifications are generally equivalent.
<sup>c</sup>) P-value was obtained from a cumulative link model while adjusting for age and sex.
<sup>d</sup>) Neurological diseases included stroke, Parkinson disease, dementia, and multiple sclerosis.
<sup>e</sup>) P-values were obtained from a logistic regression model while adjusting for age and sex.
<sup>f</sup>) P-values were obtained from multivariable linear regression model while adjusting for sex.
<sup>g</sup>) P-values were obtained from multivariable linear regression model while adjusting for age, age<sup>2</sup>, sex and ETIV.
*Abbreviations:* ETIV = estimated total intracranial volume, ISCED = International Standard Classification of Education, SD = standard deviation, TBV = total brain volume

**Table 2.**
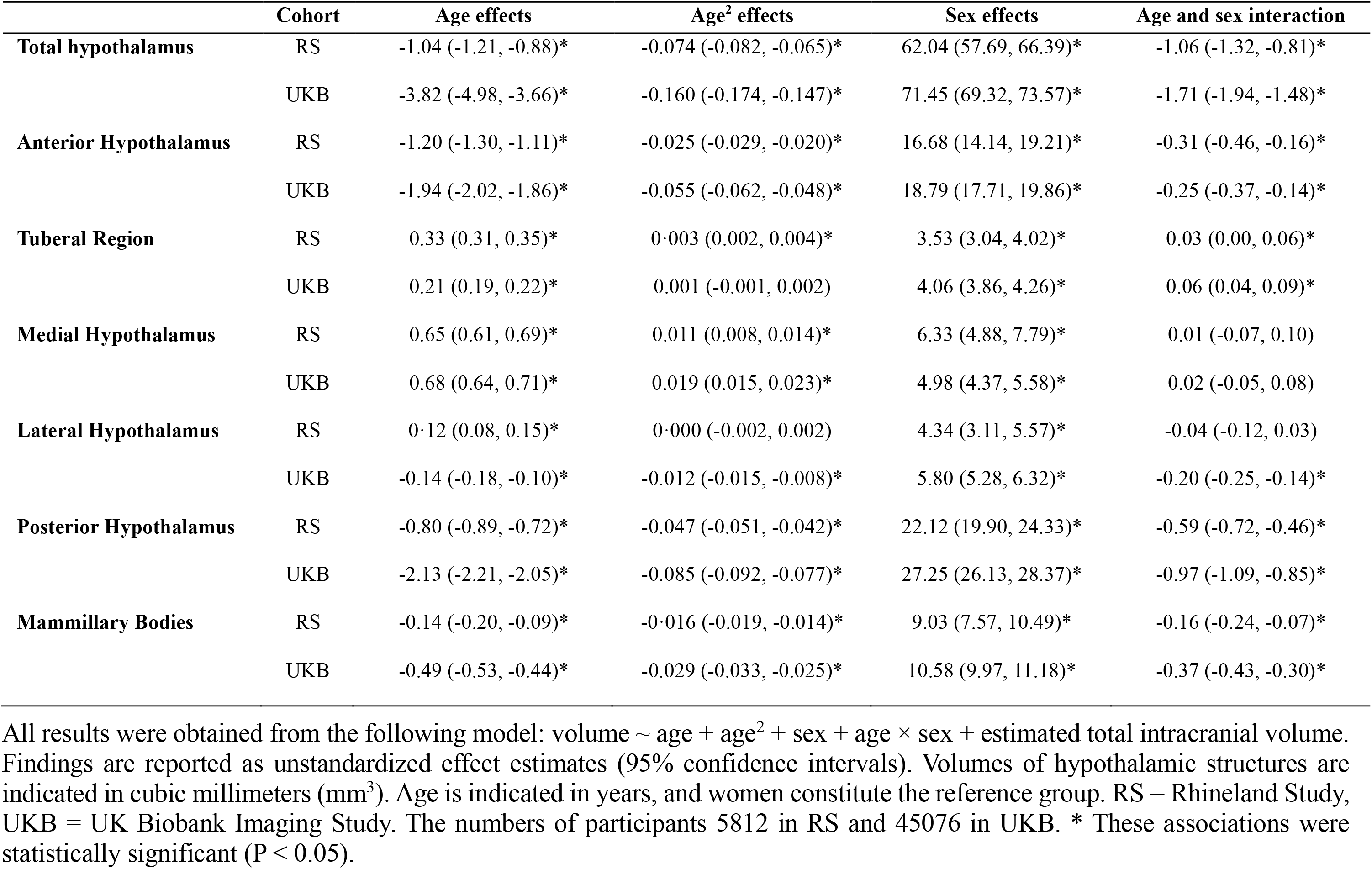
Age and sex effects on volumes of hypothalamic structures.

|  | Cohort | Age effects | Age<sup>2</sup> effects | Sex effects | Age and sex interaction |
| --- | --- | --- | --- | --- | --- |
| **Total hypothalamus** | RS | -1.04 (-1.21, -0.88)* | -0.074 (-0.082, -0.065)* | 62.04 (57.69, 66.39)* | -1.06 (-1.32, -0.81)* |
|  | UKB | -3.82 (-4.98, -3.66)* | -0.160 (-0.174, -0.147)* | 71.45 (69.32, 73.57)* | -1.71 (-1.94, -1.48)* |
| **Anterior Hypothalamus** | RS | -1.20 (-1.30, -1.11)* | -0.025 (-0.029, -0.020)* | 16.68 (14.14, 19.21)* | -0.31 (-0.46, -0.16)* |
|  | UKB | -1.94 (-2.02, -1.86)* | -0.055 (-0.062, -0.048)* | 18.79 (17.71, 19.86)* | -0.25 (-0.37, -0.14)* |
| **Tuberal Region** | RS | 0.33 (0.31, 0.35)* | 0.003 (0.002, 0.004)* | 3.53 (3.04, 4.02)* | 0.03 (0.00, 0.06)* |
|  | UKB | 0.21 (0.19, 0.22)* | 0.001 (-0.001, 0.002) | 4.06 (3.86, 4.26)* | 0.06 (0.04, 0.09)* |
| **Medial Hypothalamus** | RS | 0.65 (0.61, 0.69)* | 0.011 (0.008, 0.014)* | 6.33 (4.88, 7.79)* | 0.01 (-0.07, 0.10) |
|  | UKB | 0.68 (0.64, 0.71)* | 0.019 (0.015, 0.023)* | 4.98 (4.37, 5.58)* | 0.02 (-0.05, 0.08) |
| **Lateral Hypothalamus** | RS | 0.12 (0.08, 0.15)* | 0.000 (-0.002, 0.002) | 4.34 (3.11, 5.57)* | -0.04 (-0.12, 0.03) |
|  | UKB | -0.14 (-0.18, -0.10)* | -0.012 (-0.015, -0.008)* | 5.80 (5.28, 6.32)* | -0.20 (-0.25, -0.14)* |
| **Posterior Hypothalamus** | RS | -0.80 (-0.89, -0.72)* | -0.047 (-0.051, -0.042)* | 22.12 (19.90, 24.33)* | -0.59 (-0.72, -0.46)* |
|  | UKB | -2.13 (-2.21, -2.05)* | -0.085 (-0.092, -0.077)* | 27.25 (26.13, 28.37)* | -0.97 (-1.09, -0.85)* |
| **Mammillary Bodies** | RS | -0.14 (-0.20, -0.09)* | -0.016 (-0.019, -0.014)* | 9.03 (7.57, 10.49)* | -0.16 (-0.24, -0.07)* |
|  | UKB | -0.49 (-0.53, -0.44)* | -0.029 (-0.033, -0.025)* | 10.58 (9.97, 11.18)* | -0.37 (-0.43, -0.30)* |

**Figure 1.**
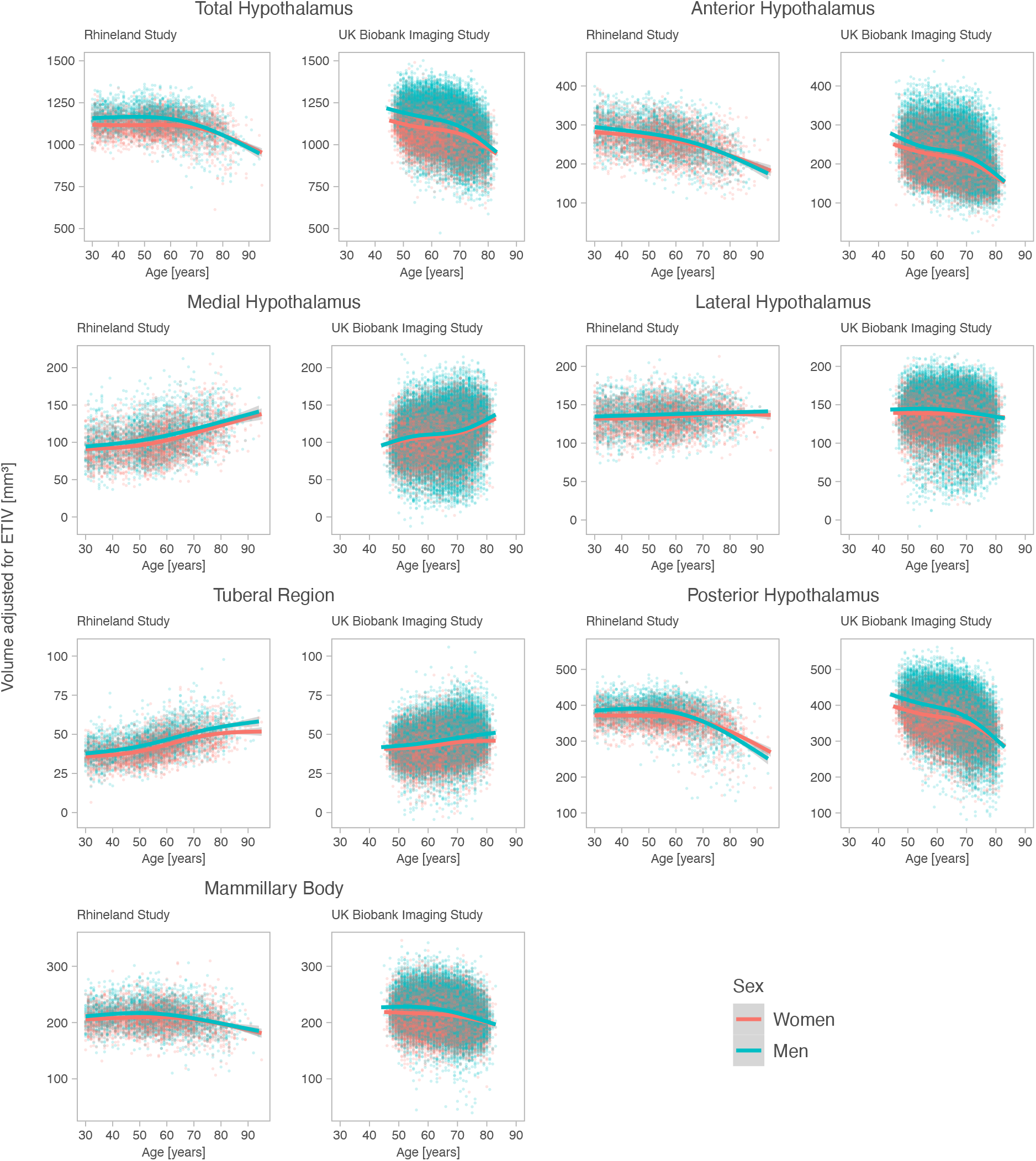
The associations of age and sex with volumes of hypothalamic structures in the Rhineland Study and the UK Biobank Imaging Study cohorts. Volumes of hypothalamic structures in cubic millimeters (mm^3^) were adjusted for estimated total intracranial volume. Plots on the left side indicate results from the Rhineland Study, while those on the right side indicate results from the UK Biobank Imaging Study. Each point represents one participant colored by sex (red for women and green for men). Overlapping points have a darker color. The trend curves are based on spline regression with four degrees of freedom.

Volumes of all seven hypothalamic structures were significantly larger in men compared to women in both RS and UKB (differences between 3.53 to 62.04 mm^3^ in RS, and between 4.06 to 71.45 mm^3^ in UKB), independent of ETIV. However, for volumes of four hypothalamic structures (TH, AH, PH, and MMB), we found that although younger men started with larger hypothalamic volumes, the volumes of these structures tended to decrease more sharply with age in men compared to women. In contrast, the difference in TBR volume between men and women became progressively larger with age (**Table 2** and **Figure 1)**. A visual summary of the effects of age and sex on volumes of different hypothalamic structures is provided in **Supplementary Figure 4**.

### Cognitive correlates

Larger total hypothalamus volumes were associated with better global cognition (β ± standard error (SE): 0.025 ± 0.017 [RS] and 0.026 ± 0.007 [UKB], both p<0.005), and total memory (0.030 ± 0.022 [RS] and 0.021 ± 0.009 [UKB], both p<0.007). Specifically in each subregion, larger PH volumes were associated with better global cognition (0.036 ± 0.014 [RS] and 0.028 ± 0.006 [UKB], both p<0.001), total memory (0.038 ± 0.018 [RS] and 0.020 ± 0.008 [UKB], both p<0.001), and executive function (0.053 ± 0.021 [RS] and 0.039 ± 0.011 [UKB], both p<0.001) (**Figure 2** and **Supplementary Table 6**). Larger MMB were associated with better total memory (0.020 ± 0.016 [RS] and 0.017 ± 0.008 [UKB], both p<0.02), episodic verbal memory (0.046 ± 0.023 [RS] and 0.011 ± 0.008 [UKB], both p<0.009), and processing speed (0.021 ± 0.018 [RS] and 0.023 ± 0.008 [UKB], both p<0.03), while larger AH volumes were related to better executive function (0.030 ± 0.018 [RS] and 0.026 ± 0.011 [UKB], both p<0.002). Conversely, a larger TBR volume was associated with worse global cognition (-0.038 ± 0.014 [RS] and -0.011 ± 0.006 [UKB], both p<0.001), executive function (-0.058 ± 0.020 [RS] and -0.020 ± 0.011 [UKB], both p<0.001), as well as processing speed (-0.026 ± 0.020 [RS] and -0.020 ± 0.007 [UKB], both p<0.02). Similarly, a larger MH volume was associated with worse global cognition (-0.013 ± 0.012 [RS] and -0.015 ± 0.006 [UKB], both p<0.05), and executive function (-0.033 ± 0.018 [RS] and -0.029 ± 0.011 [UKB], both p<0.001) (**Figure 2, Supplementary Table 6**). The summary of subregion-specific cognitive correlates is provided in **Figure 3**.

**Figure 2.**
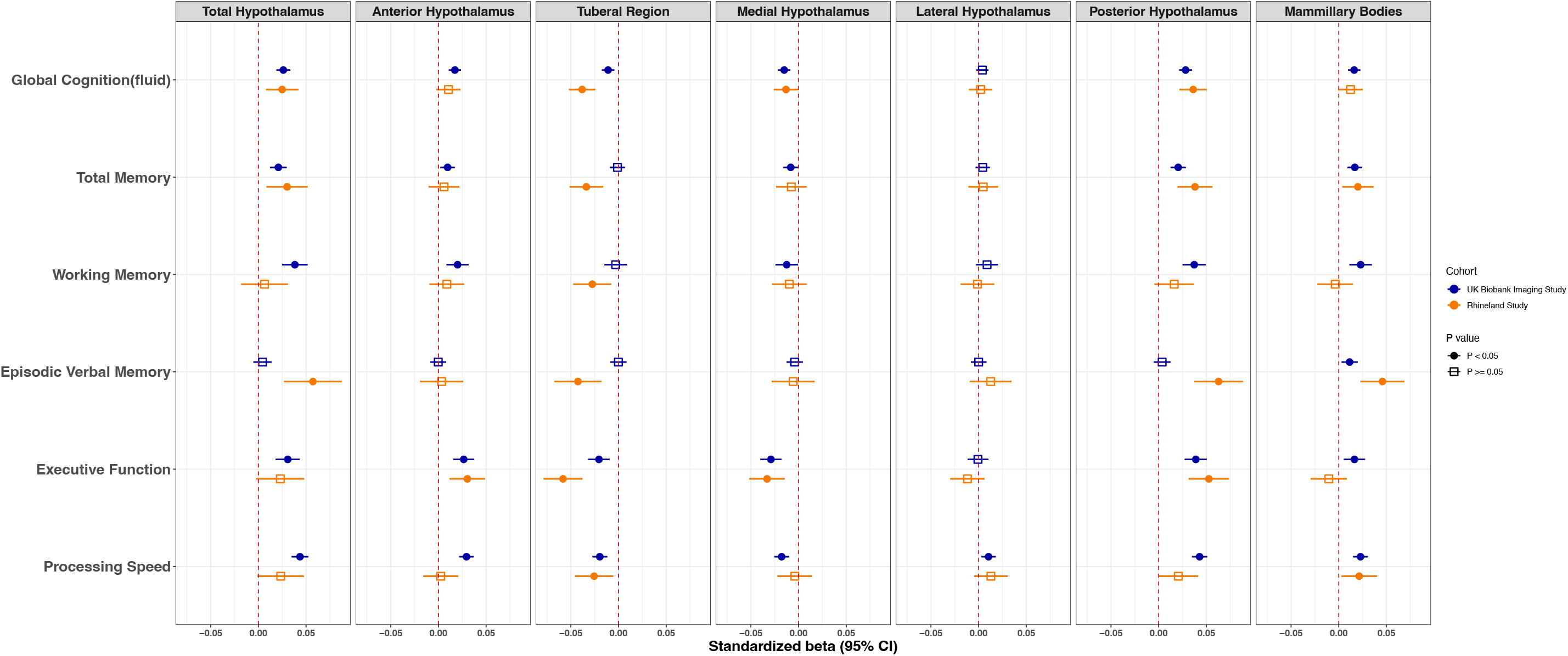
Standardized associations of volumes of hypothalamic structures with domain-specific cognitive performance in the Rhineland Study and the UK Biobank Imaging Study. The effect estimates were adjusted for age, age^2^, sex, education level and estimated total intracranial volume. The solid circles represent the statistically significant point estimates (i.e., p-value < 0.05), while the open squares represent the statistically non-significant ones.

**Figure 3.**
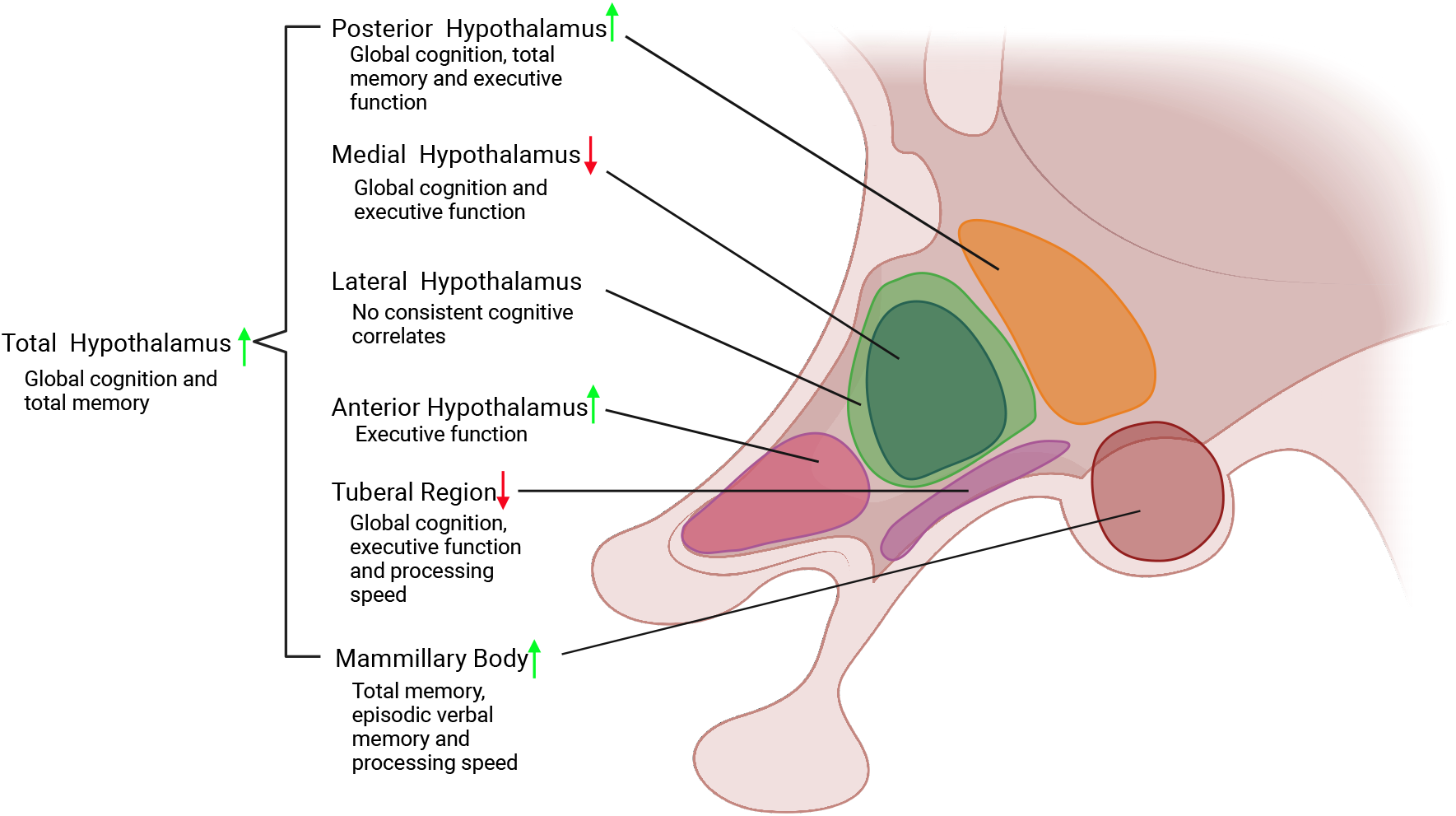
Hypothalamic structures and their cognitive correlates. The schematic figure shows the hypothalamus in a sagittal view, depicting the medial hypothalamus superimposed on the lateral part. All the statistically significant associations (i.e., p-value < 0.05 in both cohorts) between volumes of different hypothalamic structures and cognitive domain scores are shown. The green upward arrows indicate that larger volumes were associated with better cognitive performance, whereas the red downward arrows indicate that larger volumes were related to worse cognitive performance.

Despite not reaching statistical significance in both datasets, most other associations of hypothalamic structural volumes and cognitive test scores were also directionally consistent between RS and UKB (**Figure 2**). Except for LH, all hypothalamic structural volumes were significantly associated with global cognition, executive function and processing speed in UKB, which was also largely confirmed in RS. Conversely, volumes of TH, TBR, PH, and MMB were significantly associated with episodic verbal memory in RS, with only the relationship with MMB also reaching statistical significance in UKB.

### Cortisol and MH volume

There was an age-dependent association between MH volume and hair cortisol levels (p ≈ 0.020 for MH volume × age interaction effect). Age stratified analysis, based on tertiles of the age distribution, revealed that although the association between MH volume and hair cortisol levels was not significant in the younger age groups (i.e., 30-48 years and 49-61 years), larger MH volumes were related to increased cortisol levels in the oldest age group (i.e., 62-95 years; 0.053 ± 0.021, p ≈ 0.001). This effect size amounts to about 13% ± 5% increase in cortisol levels per SD increase of MH volume (**Supplementary Figure 5**).

### Sensitivity analysis

The associations of hypothalamic volumes and global cognitive function largely remained similar when the first principal component was used as a summary measure of overall cognitive function (**Supplementary Figure 6**). Similarly, exclusion of participants with known neurological diseases did not materially change the magnitude or the statistical significance of any of our findings (data not shown).

All results were obtained from the following model: volume ∼ age + age^2^ + sex + age × sex + estimated total intracranial volume. Findings are reported as unstandardized effect estimates (95% confidence intervals). Volumes of hypothalamic structures are indicated in cubic millimeters (mm^3^). Age is indicated in years, and women constitute the reference group. RS = Rhineland Study, UKB = UK Biobank Imaging Study. The numbers of participants 5812 in RS and 45076 in UKB. * These associations were statistically significant (P < 0.05).

## Discussion

We present the combined findings of two large-scale population-based studies of the effects of age and sex on the volumes of the hypothalamus and its subregions, as well as the association of these hypothalamic structures with cognitive performance in the general population. We found strong region-specific age effects on the volumes of different hypothalamic structures. Although the volumes of most hypothalamic structures decreased with age, volumes of the medial and tuberal regions of the hypothalamus significantly increased with age, a finding that we could directly functionally corroborate using hair cortisol levels in a subset of the participants. Similarly, the volumes of all hypothalamic structures exhibited substantial sex differences, even after accounting for differences in head size, most of which exhibited specific dynamic changes with age. Moreover, we discovered robust associations between the volumes of different hypothalamic subregions and domain-specific cognitive function. Importantly, our findings could readily be cross-validated between the two independent large-scale population-based cohorts, and were for the most part highly consistent between them.

In line with previous small-scale studies,^8,14^ we found that the volumes of the hypothalamus and most of its subregions decreased with age. However, unlike previous studies, which largely focused on the volume of the entire hypothalamus, our region-specific analyses revealed that the volumes of two hypothalamic regions, i.e., MH and TBR, increased with age. This novel finding could indicate potential resilience of specific hypothalamic regions to age-related neurodegeneration and brain atrophy. Indeed, previous neuropathological experiments found relatively stable neuronal counts and neuronal hypertrophy in the PVN of the hypothalamus, which is included in the MH region of our parcellations scheme, during both aging and neurodegenerative diseases.^38,39^ Similarly, the volume of the arcuate nucleus, the rodent homologue of the human infundibular nucleus and the main part of the tuberal region, remained stable during aging in rats,^40^ which also supports our findings. Given that the corticotropin releasing factor-producing neurons of the PVN constitute the origin of the hypothalamic-pituitary-adrenal axis, through which secretion of the stress hormone cortisol is centrally regulated, we assessed whether MH volume was related to hair cortisol levels. Importantly, we found an age-dependent relationship between MH volume and hair cortisol levels, indicating a stronger association between MH volume and cortisol levels with advancing age. This result parallels reports of age-associated increases in cortisol secretion,^41^ and provides direct functional validation for our observation of an increase in MH volume with age. Interestingly, hyperactivation of the hypothalamic-pituitary-adrenal axis has indeed been implicated in the pathogenesis of common age-associated neurodegenerative diseases like Alzheimer and Parkinson disease.^23^ Together, these findings thus indicate that specific hypothalamic regions may possess resilience against age-associated neurodegeneration, and might even contribute to neuropathology elsewhere in the brain.

Our findings parallel those from previous small-scale studies indicating a relatively larger volume of the hypothalamus in men compared to women, even after adjusting for ETIV.^14^ In addition, we extend previous findings by showing that these relative volumetric sex differences are not static, but exhibit substantial region-specific temporal dynamics across age. Indeed, for most hypothalamic regions the relative sex differences in volumetric measures diminished with advancing age, with the notable exception of TBR for which the volumetric sex difference increased with age. Interestingly, neuropathological experiments have found region-specific sexually dimorphic changes in both the size and number of different neuronal subpopulations of the hypothalamus, notably the supraoptic nucleus and the PVN.^42^ These sexually dimorphic changes may be attributed to age-related changes in sex hormone levels, spanning the entire period from the intrauterine to postnatal phase, adulthood, and old age.^42^ Various hypothalamic nuclei are closely involved in the regulation of the stress response, dysregulation of which has been postulated to put women at an increased risk of both mood disorders and neurodegenerative diseases, especially Alzheimer disease.^43^ Therefore, further studies aimed at delineating the sexually dimorphic role of the hypothalamus and its subregions could be instrumental in the elucidation of the neurobiological basis of sex differences in risk factors and causes of a range of (neuropsychiatric) disorders.

We discovered robust associations between the volumes of different hypothalamic subregions and domain-specific cognitive performance. Larger PH volume was consistently associated with better global cognition, total memory and executive function in both RS and UKB cohorts. Previous studies demonstrated the posterior hypothalamic area to be critical for synchronizing the theta rhythm,^44^ which could enhance cognitive performance.^45^ Moreover, this region harbors the supramammillary nucleus, which could modulate memory function through its extensive connections to the hippocampus.^2^ We also found a strong relationship between MMB volume and total memory, as well as episodic verbal memory, which extends findings form previous (animal) studies,^19,20^,46 and highlights the role of the fornix-mammillary body circuit in regulating episodic memory function.^47^ Notably, MMB volume was also related to processing speed in both datasets, which is in line with a recent clinical case-control study in children.^48^ Similarly, a larger AH volume was related to better executive function, possibly due to the localization of the suprachiasmatic and the ventral preoptic nucleus within this region. These nuclei are crucial for the regulation of circadian rhythm and sleep,^49^ which are strongly associated with better executive function.^50^ Apart from their association with age, larger volumes of the MH and TBR regions were also related to worse age-adjusted cognitive function, including global cognitive performance, executive function and processing speed. Although the precise underlying mechanisms remain to be elucidated, hyperactivation of the hypothalamic-pituitary-adrenal axis may play an important role in mediating this association as high cortisol levels have been associated with an increased risk of (age-associated) cognitive decline and neurodegenerative diseases^.23,24^ Moreover, chemogenetic activation of Agrp neurons in the arcuate nucleus of mice reduced their cognitive performance, which also points towards an alternative pathway through which increased TBR volume could be linked to worse cognitive performance.^25^ However, given the highly complex architecture of the human hypothalamus and the lack of studies assessing its role in relation to cognition, more mechanistic experiments are warranted to elucidate the underlying neurobiological pathways that could account for the region-specific associations between different hypothalamic structures and performance on various cognitive domains.

Our study has both strengths and limitations. First, although most of our results were highly consistent between the RS and UKB cohorts, there were also some exceptions. In the RS cohort, we found more significant associations between hypothalamic volumes and episodic memory, which may be due to the tests used in RS (AVLT immediate and delayed recall), which are more comprehensive compared to those employed by UKB (paired associated learning and pairs matching). Conversely, more significant volumetric associations with processing speed were observed in UKB, potentially due to the more comprehensive set of test batteries employed for the assessment of this particular cognitive domain (including the Trail-Making Test A, Reaction Time Test, and Symbol Digit Substitution Test). Second, the findings involving the LH region were inconsistent. This could be due to technical challenges in accurately delineating the lateral boundaries of the hypothalamus, which are formed by diffuse white matter tracts, on MR images. This may have been further exacerbated by differing spatial imaging resolutions in the two cohorts.^26^ Third, different cognitive tests were used in the two cohorts. To address this issue, we defined cognitive domain scores based on the nature of the tests employed. Importantly, except for the LH region, all our statistically significant results were directionally consistent between the two cohorts, supporting the robustness and generalizability of the findings. Lastly, due to the cross-sectional nature of our study, we could not assess the temporal dynamics between volumetric changes in hypothalamic structures and cognitive performance, which should be the focus of future longitudinal studies.

In conclusion, we found strong age and sex effects on volumes of different hypothalamic structures, as well as robust and region-specific associations between the volumes of these structures and performance on various cognitive domains. Thus, our findings implicate specific hypothalamic subregions as novel potential therapeutic targets against age-associated cognitive decline.

## Data Availability

The Rhineland Study's dataset is not publicly available because of data protection regulations. Access to data can be provided to scientists in accordance with the Rhineland Study's Data Use and Access Policy. Requests for further information or to access the Rhineland Study's dataset should be directed to. All individual level data from the UK Biobank Imaging Study used in the current manuscript are available through the UK Biobank Resource (https://www.ukbiobank.ac.uk). This research has been conducted using the UK Biobank Resource under Application Number 82056.

## Acknowledgments

We would like to thank the Rhineland Study team for supporting the data acquisition and management. We also would like to thank Konstantinos Melas, Annabell Coors, and Daniel Rusman for technical advice and assistance. This work was supported by DZNE institutional funds, the Federal Ministry of Education and Research of Germany (031L0206, 01GQ1801), an Alzheimer’s Association Research Grant (Award Number: AARG-19-616534), the Chan Zuckerberg Initiative (Project FastSurfer, Grant Number: EOSS5 2022-252594), the Helmholtz-AI project DeGen (ZT-I-PF-5-078), and NIH (R01 LM012719, R01 AG064027, R56 MH121426, and P41 EB030006). Peng Xu is supported by a scholarship from China Scholarship Council, and N. Ahmad Aziz is supported by a European Research Council Starting Grant (Number: 101041677). Part of this research has been conducted using the UK Biobank Resource under Application Number 82056.

