## Supplementary Tables and Figures for "Hypothalamic integrity is associated with age, sex and cognitive function across lifespan: A comparative analysis of two large population-based cohort studies"

**Supplementary table 1. Sequence parameters for the T1-weighted and T2-weighted versions in the Rhineland Study.**

| **T1w sequence** | | | **T2w sequence** | | | | |
|---|---|---|---|---|---|---|---|
| Protocol | A multi-echo magnetization prepared rapid gradient echo (MPRAGE) sequence,[1] with 2D acceleration.[2] | | Protocol | A 3D turbo-spin-echo (TSE) sequence, with variable flip angles.[3] | | | |
| | Version | | | Version | | | |
| Parameters | T1w$^{a}$ | T1w$^{b}$ | Parameters | T2w$^{a}$ | T2w$^{b}$ | T2w$^{c}$ | T2w$^{d}$ |
| Repetion time (TR) | 2560 ms | | Repetion time (TR) | 2800 ms | | | |
| Inversion time (TI) | 1100 ms | | Echo time (TE) | 405 ms | | | |
| Matrix size | 320 × 320 × 224 | | Matrix size | 320 × 320 × 224 | | | |
| Flip angle | 7° | | Phase-encoding direc.±± | A>P | R>L | A>P | A>P |
| PI acc. factor | 1×3 | 1×2 | PI acc. factor | 3×1 | | 2×1 | 1×2± |
| Echo time (TE) | 2.94 ms* | 1.68 ms to 6.51 ms** | PI ref. scan | Integrated | | External | |
| Acquisition time (TA) | 3:43 minutes | 6:35 minutes | Acquisition time (TA) | 3:57 minutes | 4:30 minutes | 4:47 minutes | |
| Readout bandwith | 240 Hz/pixel | 740 Hz pixel | | | | | |

To date, there have been two versions of the T1w sequence (T1w$^{a\text{-}b}$) and four versions of the T2w sequence (T2w$^{a\text{-}d}$) - care was taken to preserve the image contrast between versions for both sequences.

* 1 echo, ** 4 echoes combined to 1.

\+ with one CAIPIRINHA shift,[4] ++ A: anterior, P: posterior, R: right, and L: Left.

**Supplementary table 2. Criteria for segmentation of the hypothalamic substructures.**

| Structure | Bilateral* | Labelling** |
|---|---|---|
| Total hypothalamus | No | Medial border: 3rd ventricle.<br>Lateral border: lateral border of the optic tract and the other adjacent white matter tracts.[5]<br>Anterior border: lamina terminalis attached to the optic chiasm.<br>Posterior border: vanishment of the mammillary bodies on coronal sections in the rostro-caudal direction.<br>Superior border: horizontal plane through the anterior commissure and the diencephalic fissure.[6]<br>Inferior border: optic chiasm and infundibulum anteriorly, as well as boundaries with the mammillary bodies below posteriorly. |
| Anterior hypothalamus | Yes | Medial border: 3rd ventricle.<br>Lateral border: lateral border of the optic tract and the other adjacent white matter tracts.<br>Anterior border: lamina terminalis attached to the optic chiasm.<br>Posterior border: vanishment of the anterior commissure on coronal sections in the rostro-caudal direction (coinciding with the coronal plane through the posterior border of the anterior commissure and the anterior tip of the infundibulum).<br>Superior border: horizontal plane through the anterior commissure.<br>Inferior border: optic chiasm and infundibulum. |
| Tuberal region | No | The area was defined as the region underneath the 3rd ventricle and enclosed by the mammillary bodies caudally and the anterior hypothalamus rostrally, with its superior and inferior borders on each side defined by the horizontal planes going through the superior border of the floor of the third ventricle and the interpeduncular cistern, respectively. |
| Medial hypothalamus | Yes | Medial border: the 3rd ventricle<br>Lateral border: fornices<br>Anterior border: vanishment of the anterior commissure on coronal sections in the rostro-caudal direction.<br>Posterior border: appearance of the mammillary bodies on coronal sections in the rostro-caudal direction.<br>Superior border: the diencephalic fissure.<br>Inferior border: the boundaries of the tuberal region underneath.[7] |
| Lateral hypothalamus | Yes | Medial border: fornices.<br>Lateral border: optic tract and the other adjacent white matter tracts.<br>Anterior border: vanishment of the anterior commissure on coronal sections in the rostro-caudal direction.<br>Posterior border: appearance of the mammillary bodies on coronal sections in the rostro-caudal direction.<br>Superior border: the diencephalic fissure.<br>Inferior border: the boundaries of the tuberal region and basal cistern underneath<br>. |
| Posterior hypothalamus | Yes | Medial border: the 3rd ventricle.<br>Lateral border: white matter tracts.<br>Anterior border: appearance of the mammillary bodies on coronal sections in the rostro-caudal direction.<br>Posterior border: vanishment of the mammillary bodies on coronal sections in the rostro-caudal direction.<br>Superior border: horizontal plane through the diencephalic fissure.<br>Inferior border: boundaries with the mammillary bodies below. |
| Mammillary bodies | Yes | Two small, rounded structures at the caudal end of the 3rd ventricle. These structures were labelled using both coronal sections in the rostro-caudal direction and axial sections in the dorso-medial direction on T1 weighted images. |

*) Bilateral structures were defined as those regions that could be separated into a (non-contiguous) left and right half with respect to the midsagittal plane.
**) Labelling was mainly done using T1 weighted images, unless specified otherwise.

**Supplementary table 3. Cognitive assessments in the two cohorts.**

| | Test | Outcome | Mean (SD) | Procedure* |
|---|---|---|---|---|
| **Rhineland Study** | | | | |
| Working memory | Corsi Block-tapping span forward | Maximum number of blocks being correctly remembered in forward order | 4.9 (1.1) | Z-score |
| | Corsi Block-tapping span backward | Maximum number of blocks being correctly remembered in backward order | 4.8 (1.0) | Z-score |
| | Digit span forward | Maximum number of digits being correctly remembered in forward order | 6.4 (1.2) | Z-score |
| | Digit span backward | Maximum number of digits being correctly remembered in backward order | 4.8 (1.2) | Z-score |
| Episodic verbal memory | AVLT immediate recall | Sum of correctly recalled items in recalls 1-5 | 51.6 (9.8) | Z-score |
| | AVLT time delayed recall | Number of correctly recalled items in recall 7 | 10.4 (3.2) | Z-score |
| Executive function | Word fluency | Number of distinct animals named in one minute | 25.5 (6.8) | Z-score |
| | Trail-making test B | Time to completion in seconds | 53.6 (36.2) | 1. Log 10 transforming<br>2. Reversing<br>3. Z-score |
| | Anti-saccade task | Error rate: percentage of direction errors | 31.5 (23.5) | 1. Reversing<br>2. Z-score |
| Processing speed | Trail-making test A | Time to completion in seconds | 36.9 (17.8) | 1. Log 10 transforming<br>2. Reversing<br>3. Z-score |
| | Pro-saccade task | Mean saccadic latency in correct trials in milliseconds | 189.6 (28.0) | 1. Reversing<br>2. Z-score |

| **UK Biobank Imaging Study** | | | | |
|---|---|---|---|---|
| Working memory | Numeric memory | Maximum digits remembered correctly | 6.7 (1.3) | Z-score |
| Episodic verbal memory | Pairs matching | Number of incorrect matches in round 2 | 3.6 (2.9) | 1. Log 10 transforming<br>2. Reversing<br>3. Z-score |
| | Paired associated learning | Number of word pairs correctly associated | 6.8 (2.7) | 1. Log 10 transforming<br>2. Z-score |
| Executive function | Trail making test-B | Time to completion in seconds | 57.7 (25.3) | 1. Log 10 transforming<br>2. Reversing<br>3. Z-score |
| Processing speed | Reaction time | Mean time to correctly identify matches in milliseconds | 597.2(110.6) | 1. Log 10 transforming<br>2. Reversing<br>3. Z-score |
| | Trail making test-A | Time to completion in seconds | 22.8 (8.7) | 1. Log 10 transforming<br>2. Reversing<br>3. Z-score |
| | Symbol digit substitution | Number of symbol digit matches made correctly | 18.8 (5.3) | Z-score |

[*]) Procedure refers to the statistical method used to process the raw data into the final variables for further analysis.
Abbreviation: AVLT = The (Rey) Auditory Verbal Learning and Memory Test.

**Supplemental Table 4. Comparison between analyzed sample and excluded participants in RS**

| | Analysed sample<br>N = 5812 | Excluded participants<br>N = 2506 | P-value |
|---|---|---|---|
| Age, years | 55.2 (13.6) | 57.4 (14.3) | <0.001[a] |
| Sex | | | <0.001[a] |
| Women | 3,362 (58%) | 1,332 (53%) | |
| Men | 2,450 (42%) | 1,174 (47%) | |
| Education level[b] | | | <0.002[c] |
| High | 3,793 (66%) | 1,529 (62%) | |
| Middle | 1,869 (32%) | 861 (35%) | |
| Low | 95 (1.7%) | 67 (2.7%) | |
| Neurological diseases | 113 (1.9%) | 99 (4.0%) | <0.001[e] |

Data are shown as mean (standard deviation) or n (%) for continuous and categorical variables, respectively.
[a]) P-value were obtained from Welch's Two Sample t-test for continuous variables and from Pearson's Chi-squared test for categorical variables.
[b]) Education level was defined as follows: High (ISCED-11 levels 6-8), Middle (ISCED-11 levels 3-5), and Low (ISCED-11 levels 0-2).
[c]) P-value was obtained from a cumulative link model while adjusting for age and sex.
[d]) Neurological diseases included stroke, Parkinson disease, dementia, and multiple sclerosis.
[e]) P-values were obtained from a logistic regression model while adjusting for age and sex.
Abbreviation: N = number of participants.

**Supplemental Table 5. Comparison between analyzed sample and excluded participants in UKB**

| | Analysed sample<br>N = 45076 | Excluded participants<br>N = 4229 | p-value |
|---|---|---|---|
| Age, years | 64.2 (7.7) | 66.5 (8.0) | <0.001[a] |
| Sex | | | <0.001[a] |
| Female | 23,950 (53%) | 1,692 (40%) | |
| Male | 21,126 (47%) | 2,537 (60%) | |
| Education level[b] | | | <0.002[c] |
| High | 21,811 (49%) | 1,921 (46%) | |
| Middle | 19,935 (45%) | 1,892 (45%) | |
| Low | 2,855 (6.4%) | 356 (8.5%) | |
| Neurological diseases[d] | 746 (1.7%) | 115 (2.7%) | <0.001[e] |

Data are shown as mean (SD) or n (%) for continuous and categorical variables, respectively.

[a]) P-value were obtained from Welch's Two Sample t-test for continuous variables and from Pearson's Chi-squared test for categorical variables.

[b]) Education level was defined as follows: High (equal to or higher than college level, Middle (secondary education level), and Low (below secondary level).

[c]) P-value was obtained from a cumulative link model while adjusting for age and sex.

[d]) Neurological diseases included stroke, Parkinson disease, dementia, and multiple sclerosis.

[e]) P-values were obtained from a logistic regression model while adjusting for age and sex.

Abbreviation: N = number of participants.

**Supplemental Table 6. Association between cognitive domain scores and hypothalamic structures.**

| | | Total Hypothalamus | | Anterior Hypothalamus | | Tuberal Region | | Medial Hypothalamus | | Lateral Hypothalamus | | Posterior Hypothalamus | | Mammillary Bodies | |
|---|---|---|---|---|---|---|---|---|---|---|---|---|---|---|---|
| | | **β (95% CI)** | **P** | **β (95% CI)** | **P** | **β (95% CI)** | **P** | **β (95% CI)** | **P** | **β (95% CI)** | **P** | **β (95% CI)** | **P** | **β (95% CI)** | **P** |
| GC | RS N=5708 | 0.025 (0.008, 0.042) | 0.004* | 0.011 (-0.002, 0.023) | 0.10 | -0.038 (-0.052, -0.024) | <0.001* | -0.013 (-0.026, -0.001) | 0.04* | 0.002 (-0.010, 0.014) | 0.73 | 0.036 (0.022, 0.050) | <0.001* | 0.012 (-0.001, 0.025) | 0.06 |
| | UKB N=29600 | 0.026 (0.019, 0.033) | <0.001* | 0.017 (0.011, 0.024) | <0.001* | -0.011 (-0.017, -0.004) | <0.001* | -0.015 (-0.022, -0.009) | <0.001* | 0.004 (-0.002, 0.010) | 0.22 | 0.028 (0.022, 0.035) | <0.001* | 0.016 (0.010, 0.023) | <0.001* |
| TM | RS N=5726 | 0.030 (0.008, 0.052) | 0.006* | 0.006 (-0.010, 0.022) | 0.48 | -0.034 (-0.052, -0.016) | <0.001* | -0.008 (-0.024, 0.008) | 0.35 | 0.005 (-0.011, 0.020) | 0.54 | 0.038(0.020, 0.056) | <0.001* | 0.020 (0.004, 0.036) | 0.016* |
| | UKB N=30746 | 0.021 (0.012, 0.030) | <0.001* | 0.010 (-0.002, 0.017) | 0.013* | -0.001 (-0.009, 0.007) | 0.79 | -0.008 (-0.016, -0.001) | 0.04* | 0.004 (-0.003, 0.012) | 0.26 | 0.020 (0.012, 0.028) | <0.001* | 0.017 (0.009, 0.025) | <0.001* |
| WM | RS N=5732 | 0.006 (-0.018, 0.031) | 0.61 | 0.009 (-0.009, 0.027) | 0.34 | -0.027 (-0.047, -0.008) | 0.007* | -0.010 (-0.028, 0.008) | 0.30 | -0.001 (-0.019, 0.016) | 0.89 | 0.016 (-0.004, 0.037) | 0.12 | -0.004 (-0.022, 0.013) | 0.69 |
| | UKB N=31444 | 0.038 (0.025, 0.052) | <0.001* | 0.020 (0.008, 0.032) | <0.001* | -0.003 (-0.015, 0.009) | 0.63 | -0.012 (-0.024, -0.001) | 0.04* | 0.009 (-0.003, 0.020) | 0.13 | 0.037 (0.025, 0.049) | <0.001* | 0.030 (0.011, 0.035) | <0.001* |
| EVM | RS N=5748 | 0.057 (0.027, 0.087) | <0.001* | 0.003 (-0.019, 0.026) | 0.78 | -0.042 (-0.067, -0.018) | <0.001* | -0.006 (-0.028, 0.017) | 0.62 | 0.013 (-0.009, 0.034) | 0.26 | 0.063 (0.037, 0.088) | <0.001* | 0.046 (0.023, 0.069) | <0.001* |
| | UKB N=31151 | -0.004 (-0.005, 0.014) | 0.37 | 0.000 (-0.009, 0.008) | 0.96 | 0.000 (-0.009, 0.008) | 0.98 | -0.004 (-0.012, 0.004) | 0.35 | 0.000 (-0.008, 0.008) | 0.99 | 0.004 (-0.005, 0.012) | 0.42 | 0.011 (0.003, 0.020) | 0.008* |

| | | | | | | | | | | | | | |
|---|---|---|---|---|---|---|---|---|---|---|---|---|---|
| EF | RS N=5728 | 0.023 (-0.002, 0.048) | 0.07 | 0.030 (0.012, 0.049) | 0.001* | -0.058 (-0.078, -0.038) | <0.001* | -0.033 (-0.052, -0.015) | <0.001* | -0.012 (-0.030, 0.006) | 0.20 | 0.053 (0.031, 0.074) | <0.001* | -0.010 (-0.029, 0.008) | 0.28 |
| | UKB N=30023 | 0.031 (0.018, 0.043) | <0.001* | 0.026 (0.015, 0.038) | <0.001* | -0.020 (-0.032, -0.009) | <0.001* | -0.029 (-0.040, -0.018) | <0.001* | -0.001 (-0.012, 0.010) | 0.90 | 0.039 (0.027, 0.050) | <0.001* | 0.016 (0.005, 0.028) | 0.004* |
| PS | RS N=5743 | 0.023 (-0.001, 0.048) | 0.06 | 0.002 (-0.016, 0.021) | 0.80 | -0.026 (-0.046, -0.006) | 0.012* | -0.004 (-0.022, 0.014) | 0.67 | 0.013 (-0.005, 0.031) | 0.15 | 0.021 (0.000, 0.041) | 0.005 | 0.021 (0.003, 0.040) | 0.02* |
| | UKB N=30549 | 0.043 (0.035, 0.052) | <0.001* | 0.029 (0.022, 0.037) | <0.001* | -0.020 (-0.027, -0.012) | <0.001* | -0.018 (-0.026, -0.010) | <0.001* | 0.010 (0.003, 0.018) | 0.007* | 0.043 (0.035, 0.051) | <0.001* | 0.023 (0.015, 0.031) | <0.001* |

N = number of participants with valid domain score, β = standardized estimate, 95% CI = 95% confidence interval. Abbreviations: GC = global cognition, TM = total memory, WM = working memory, EVM = episodic verbal memory, EF = executive function, PS = processing speed, RS = Rhineland Study, UKB = UK Biobank Imaging Study. All the coefficients were obtained from the following model: cognitive domain score ~ volume + age + $age^2$ + sex + education + ETIV. * The cognitive domain score was statistically significantly associated with volume ($p < 0.05$).

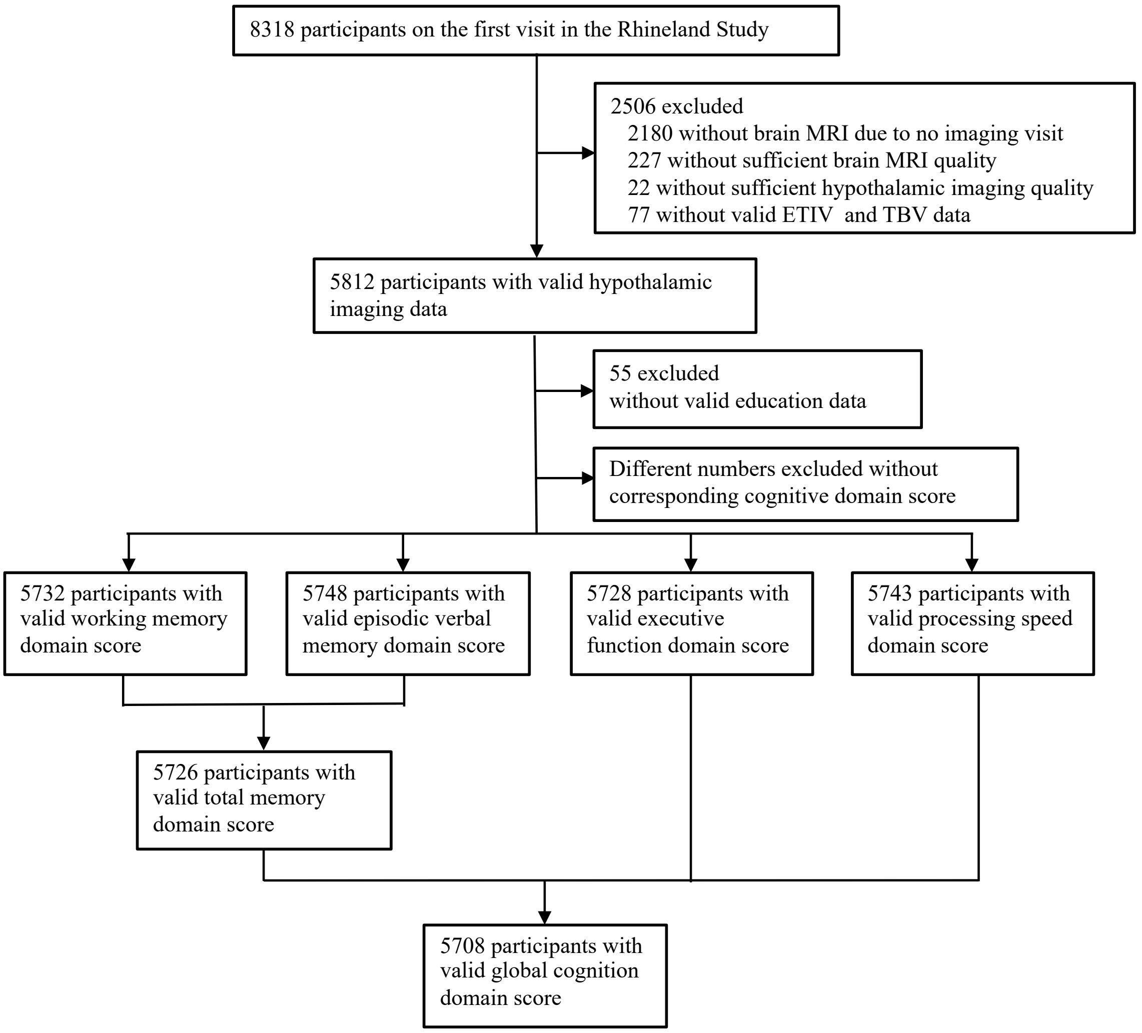

**Supplementary Figure 1.** Selection flow chart for the Rhineland Study. MRI = magnetic resonance imaging; ETIV = estimated total intracranial volume; TBV = total brain volume.

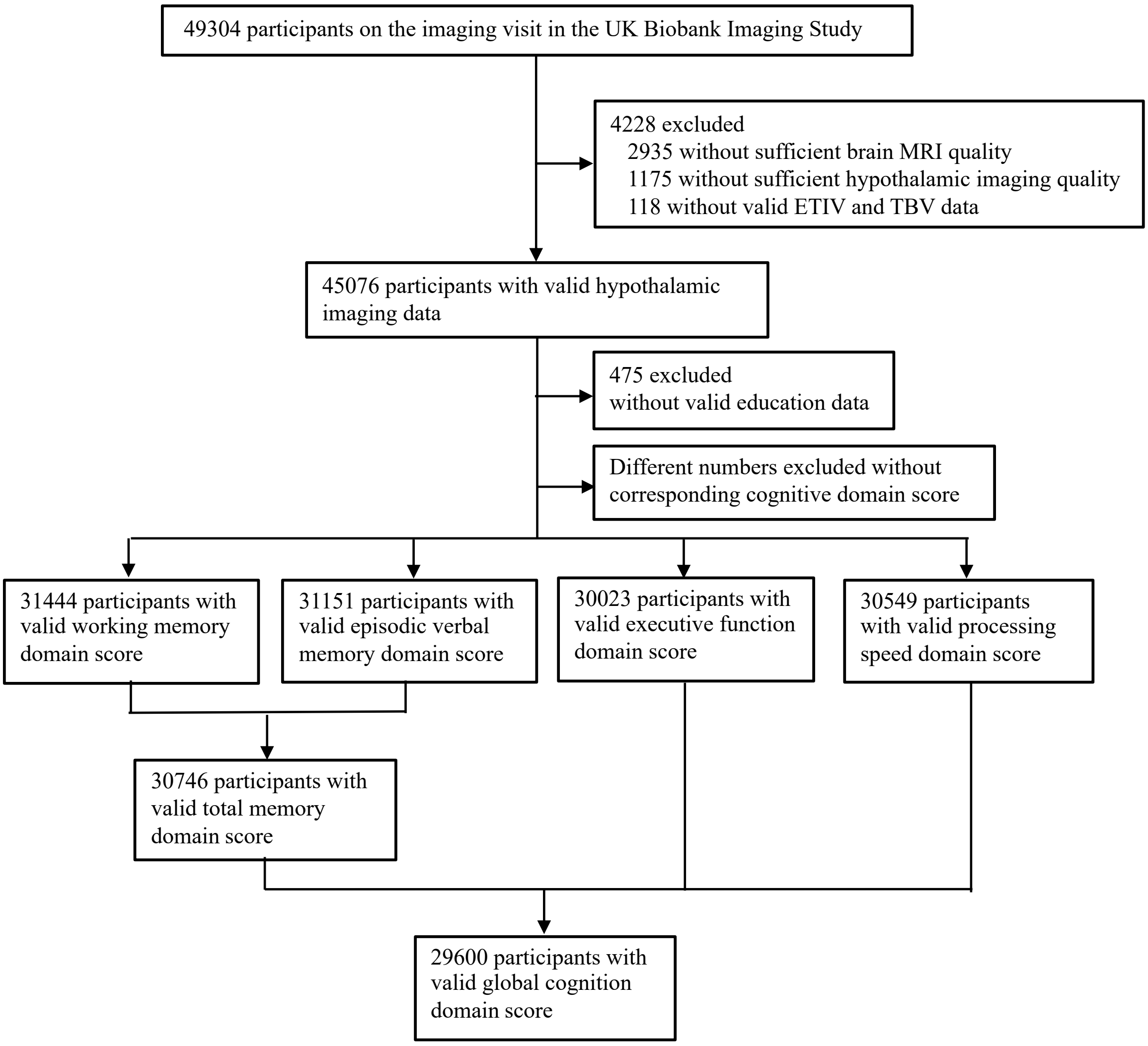

**Supplementary Figure 2.** Selection flow chart for the UK Biobank Imaging Study. MRI = magnetic resonance imaging; ETIV = estimated total intracranial volume; TBV = total brain volume.

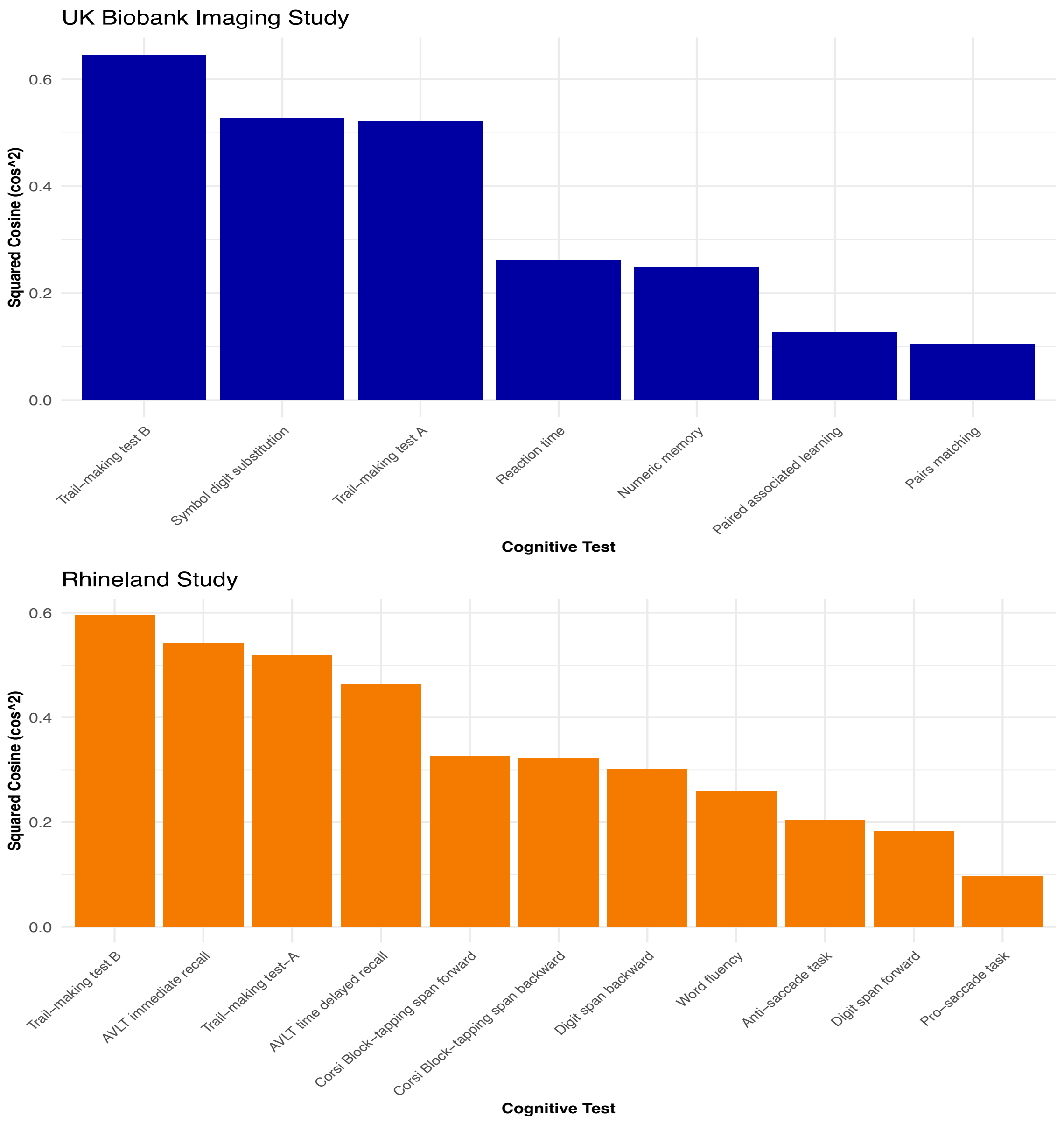

**Supplementary Figure 3. Associations between cognitive tests and squared cosine values in two cohorts.** The height of each column represents the quality of representation of each cognitive test on the first principal component.

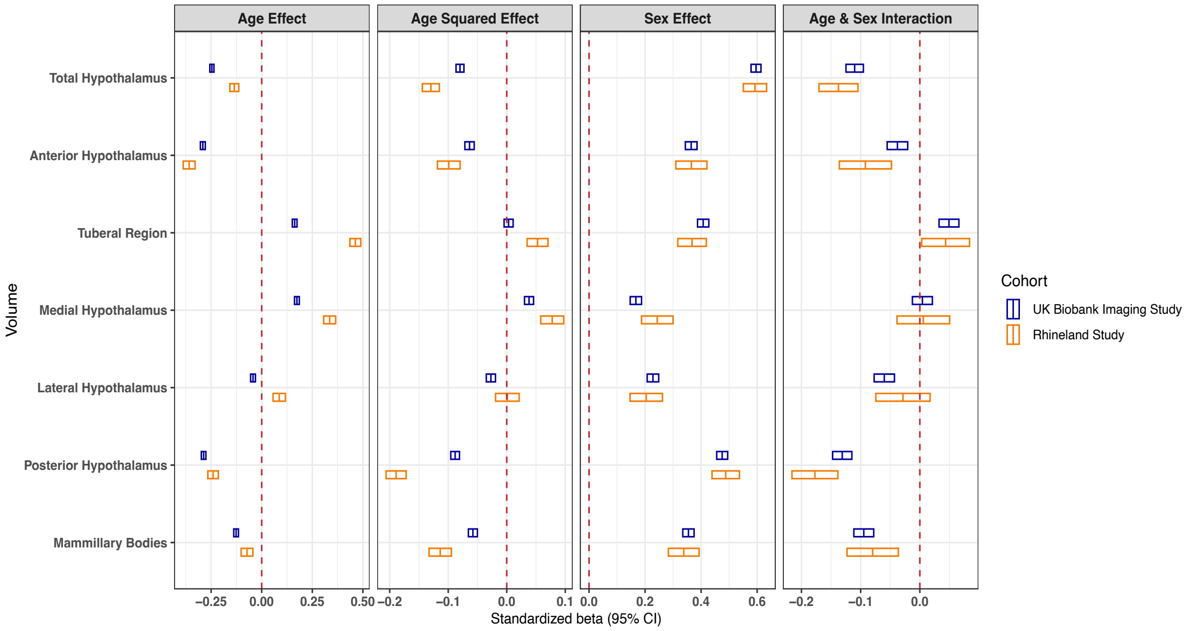

**Supplementary Figure 4. Associations of age and sex with volumes of hypothalamic structures in the two cohorts.** Each small box corresponds to a separate hypothalamic structure. Within the small boxes, the vertical lines represent the mean effect size, while the horizontal lines extending to both sides represent the associated confidence intervals. Model: volume ~ age + $age^2$ + sex + age × sex + ETIV.

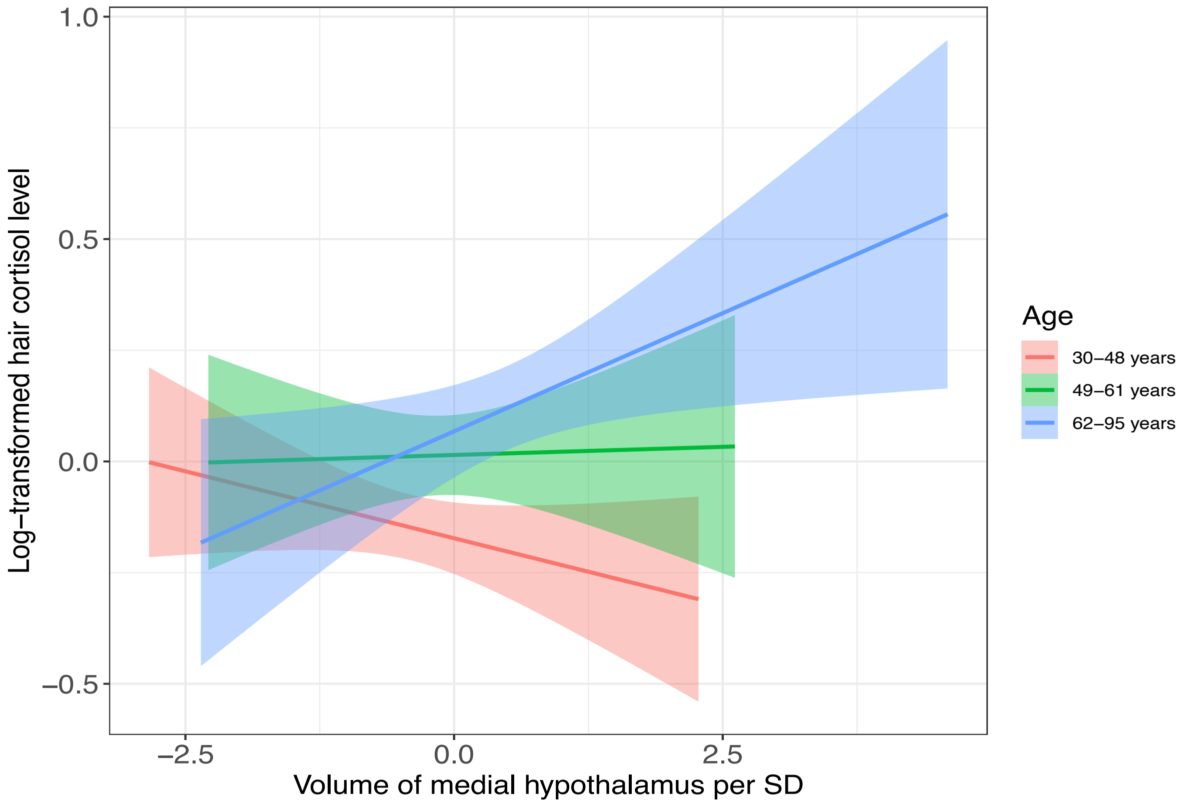

**Supplementary Figure 5. Association between volume of medial hypothalamus and hair cortisol levels stratified by age.** Lines represent the linear association between the volume of medial hypothalamus (adjusted for ETIV) and log-transformed hair cortisol levels for each tertile of age. The shaded areas represent the accompanying 95% confidence intervals of the mean.

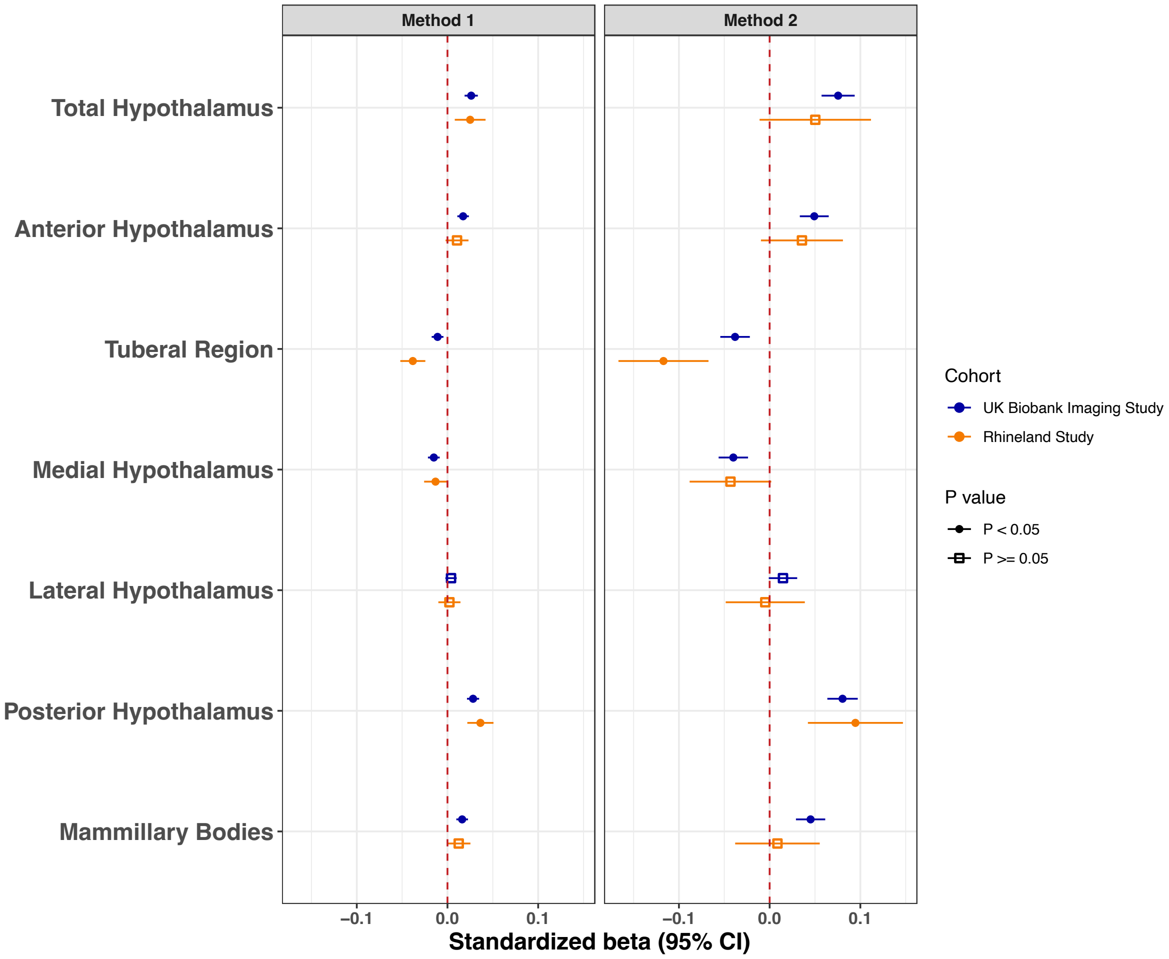

**Supplementary Figure 6. Association between global cognition and hypothalamic structures using two different methods.**
**Method 1:** Global cognition score was defined as the average of all domain-specific Z-scores.
**Method 2:** Global cognition was defined as the first principal component of the Z-scores across all tests.
The solid circles represent the statistically significant point estimates of the effect size, while the open squares represent the statistically non-significant ones.
Model: Global cognition score ~ volume + age + age$^2$ + sex + education + ETIV.
